# Development of a screening tool for pelvic floor dysfunction in female athletes: protocol of a Delphi consensus

**DOI:** 10.1101/2022.01.12.22269000

**Authors:** S Giagio, A Turolla, T Innocenti, S Salvioli, G Gava, P Pillastrini, M Vecchiato, JG Dakic, HC Frawley, K Bø

## Abstract

**Background/aim:** Several epidemiological studies have found a high prevalence of Pelvic Floor Dysfunction (PFD) among female athletes. However, according to several authors, these data could even be underestimated, both in research and clinical practice. Screening for potential PFD is often delayed and risk factors are not often evaluated. As a consequence, withdrawal from sport, negative influence on performance, worsening symptoms and unrecognized diagnosis may occur.

The aim of our research is to develop a screening tool for pelvic floor dysfunction in female athletes useful for clinicians (musculoskeletal/sport physiotherapists, sports medicine physicians, team physicians) to guide referral to a PFD expert (e.g. pelvic floor/women’s health physiotherapist, gynecologist, uro-gynecologist, urologist).

**Methods:** A 2-round modified Delphi study will be conducted to ascertain expert opinion on which combination of variables and risk factors should be included in the screening tool.

**Conclusion:** The implementation of the present screening tool into clinical practice may facilitate the referral to a PFD expert for further assessment of the pelvic floor and therefore, to identify potential dysfunction and, eventually, the related treatment pathway.

## INTRODUCTION

Pelvic floor dysfunction (PFD) refers to a group of symptoms, signs, and conditions primarily affecting women, with or without moderate-to-severe impairment of the pelvic floor muscles^1–3^.

Recently, a scoping review reported a wide range of published studies providing epidemiological data about different PFD in athletes practicing various sports^4^, highlighting that almost 90% of the literature focused only on females. Compared with non-athlete control women, athletes have a higher risk of developing urinary incontinence (UI)^5^ and also a greater prevalence rate of UI, ranging from 0% to 80% in trampolinists^6^. Regarding other PFD such as pelvic organ prolapse (POP) and anal incontinence (AI) evidence are still scant^4^. These disorders could interfere not only with the athletes ‘personal and social lives, but also could affect their performance^6–8^. Nevertheless, several authors have already discussed that it is reasonable to assume that the overall problem could be underestimated^4,9^. In fact, PFD in female athletes may be an under-recognized, under-treated, and under-researched problem^10^.

These conditions could be under-recognized for several reasons. Studies showed that the athletes’ knowledge of the pelvic floor muscles was low^8,11^, few had discussed their condition with coaches or medical personnel^7,8^, and only a minority of professionals were aware and consistently screened for PFD^12^.

In more details, findings from a cross-sectional online survey in Australia^12^ were presented at the 2021 International Continence Society (ICS) Congress. In this study, representing the only one on the topic, authors found that one in two health and exercise professionals never screened exercising women for pelvic floor symptoms, and 42.4% of healthcare professionals reported waiting for patients to disclose the condition first. Nearly 90% of professionals were willing to include screening in the future. On the other hand, 30.4% did not screen because pelvic floor questions were not included in existing screening tools or because they were not aware of which questions to ask.

Literature searches did not reveal any tool or instrument developed for musculoskeletal/sport physiotherapists and sports medicine physicians including PFD to be used in clinical practice during the assessment or medical visits. These healthcare professionals, who traditionally see and treat athletes mostly for other medical reasons, are usually not specialized in pelvic floor health, but they may play an important role in future pelvic floor health care for athletes.

In particular, the implementation of a screening tool may facilitate the referral pathway to a PFD expert (e.g. pelvic floor/women’s health physiotherapist, gynecologist, uro-gynecologist, urologist) for further assessment of the pelvic floor, and, therefore, to recognize potential dysfunction and related management.

For the initial management of some PFD, such as UI, the 6th International Consultation on Incontinence^13^ suggests that lifestyle interventions and pelvic floor muscle training (PFMT) are the first line treatment with level 1A evidence/recommendation showing that, when identified, the condition could be significantly improved.

In the framework mentioned above, a Delphi consensus could represent a first step in development of a validated screening tool.

The aim of this study is to develop a practical screening tool for PFD in female athletes useful for clinicians (musculoskeletal/sport physiotherapist, sports medicine physicians) to guide specialist referral.

## METHODS

An international research team will work on the development of the present screening tool using a Delphi modified consensus through a web-based survey.

For the reporting of the survey, the Checklist for Reporting Results of Internet E-Surveys (CHERRIES) guidelines^14^ will be used.

The research team will include 10 researchers and/or clinicians: Silvia Giagio, Andrea Turolla, Tiziano Innocenti, Stefano Salvioli, Paolo Pillastrini, Giulia Gava, Marco Vecchiato, Jodie Dakic, Helena Frawley and Kari Bø. The committee expertise will include the following: sports medicine, musculoskeletal/sport physiotherapy, pelvic floor physiotherapy, epidemiology, urogynecology.

At University of Bologna, this type of study is exempt from ethical approval of the University Ethics Committee.

### Definitions

#### Population of interest

The development of this screening tool will be addressed specifically to the female athletes of any age, of any performance level, practicing any type of sport.

Regarding the term “athlete”, the International Olympic Committee (IOC) suggests defining an elite athlete as *“Those competing at professional, Olympic or collegiate levels”*^15^. However, given the aim of the present study, this definition could be integrated with the more inclusive one proposed in 2016 by Araujo and Scharhag^16^. Authors summarized the American and European Societies of Cardiology^17–19^ definitions and suggested the minimal criteria to be defined as an active athlete as follows: *“One who are training in sports aiming to improve his/her performance, who is actively participating in sport competition, who is formally registered in a local, regional or national sport federation, and who have sport training and competition as his/her major activity (way of living) or focus of personal interest, devoting several hours in all or most of the days for these activities, exceeding the time allocated to other types of professional leisure activities”.*

The latter definition and criteria will be used along the development of the present Delphi consensus. *Condition: PFD*. For the development of the screening tool, we will refer to any type of PFD including the most common UI, POP, AI, pelvic pain and fecal incontinence^1–3^. The choice to consider any type of PFD was made mainly of two reasons: 1) the heterogeneity of epidemiological studies among the female athletes^4^, and 2) the aim of the tool. In fact, it will not be a diagnostic tool, but it will be a general instrument useful to guide for a specialist referral.

#### Target users

We chose to create a screening tool useful for clinical practice. Therefore, as target users, we identified healthcare professionals who are in close contact with athletes. In most cases, these professionals are musculoskeletal/sport physiotherapists and sports medicine physicians (including team physician); none of them is usually trained and specialized in pelvic floor health. In addition, normally athletes refer to them for other medical reasons than PFD.

A summary including essential information concerning the development of the screening tool is reported in Table 1 (**TABLE 1**).

**Table 1.**
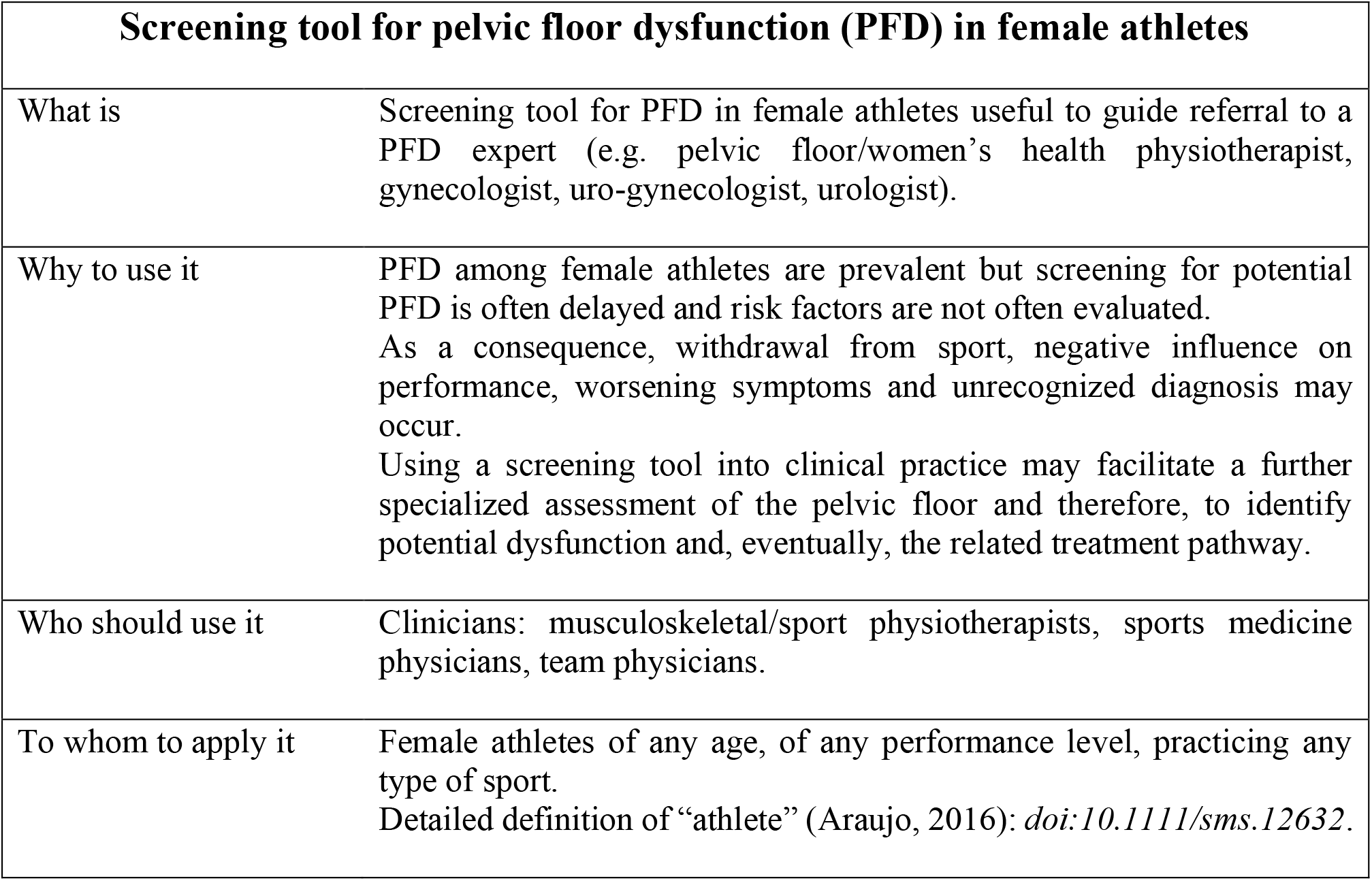
Summary table: essential information for the development of the screening tool for referral.

| **Screening tool for pelvic floor dysfunction (PFD) in female athletes** |  |
| --- | --- |
| What is | Screening tool for PFD in female athletes useful to guide referral to a PFD expert (e.g. pelvic floor/women's health physiotherapist, gynecologist, uro-gynecologist, urologist). |
| Why to use it | PFD among female athletes are prevalent but screening for potential PFD is often delayed and risk factors are not often evaluated.<br>As a consequence, withdrawal from sport, negative influence on performance, worsening symptoms and unrecognized diagnosis may occur.<br>Using a screening tool into clinical practice may facilitate a further specialized assessment of the pelvic floor and therefore, to identify potential dysfunction and, eventually, the related treatment pathway. |
| Who should use it | Clinicians: musculoskeletal/sport physiotherapists, sports medicine physicians, team physicians. |
| To whom to apply it | Female athletes of any age, of any performance level, practicing any type of sport.<br>Detailed definition of "athlete" (Araujo, 2016): *doi:10.1111/sms.12632*. |

### Identification of potential variables to include in the screening tool

In addition to the risk factors associated with PFD in women of the general population^3,20–22^, the research team will conduct a comprehensive search in MEDLINE to identify published studies that reported specific variables and risk factors for the female athletes. These variables will be presented as *items* in the survey. The search strategy is included in Table 2 (**TABLE 2**).

**Table 2.**
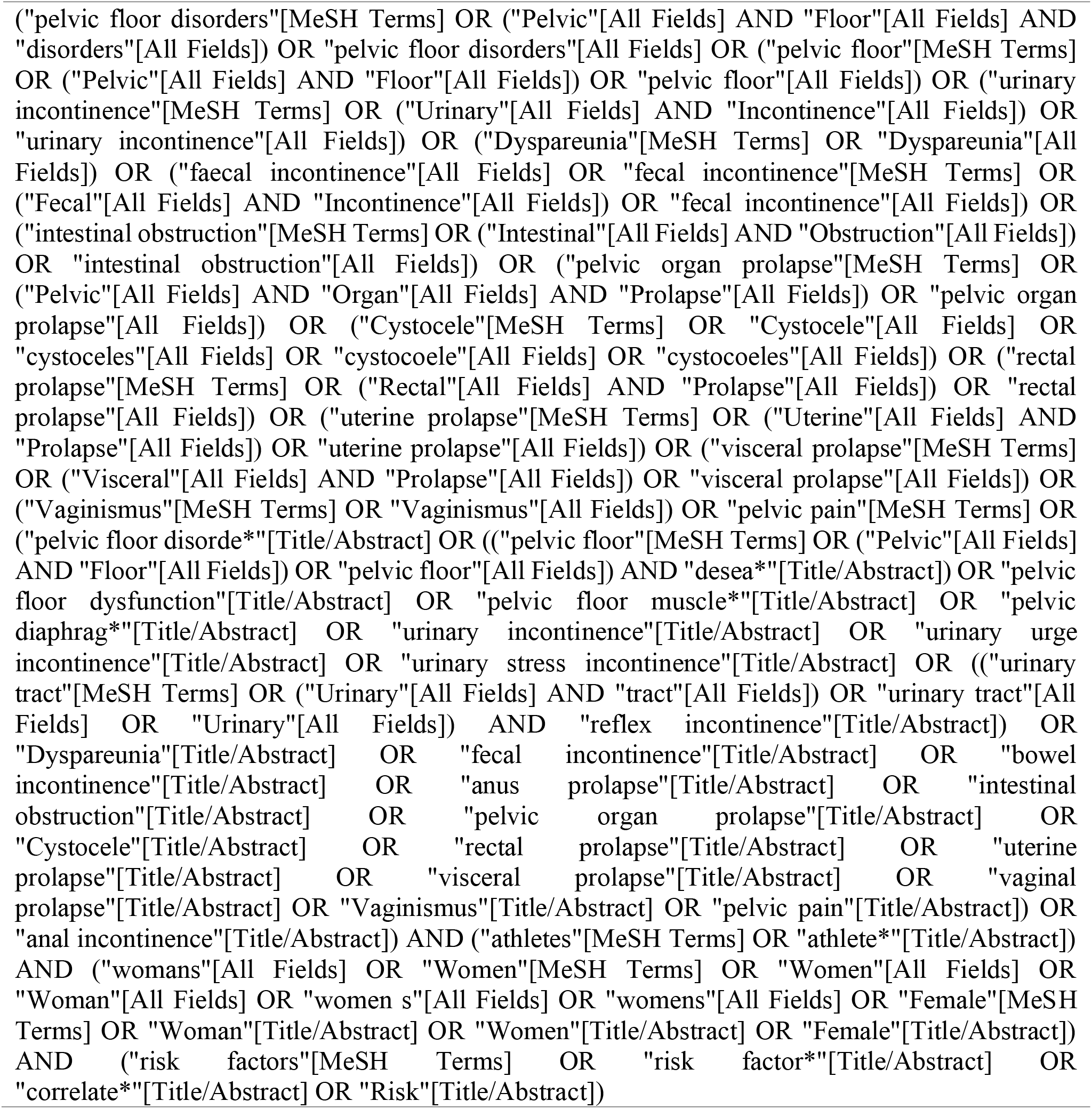
Search strategy for MEDLINE: initial identification of variables and risk factors for the female athlete to include in the screening tool. No date and language restrictions.

### Delphi study

An online modified Delphi consensus was chosen as it is a commonly used method to establish an agreement on various health- and research-related issues^23^. This is a group facilitation technique that seeks to obtain consensus on the opinions of a broad, international, and multi-stakeholder panel of ‘experts’; created on responses from others, preserving anonymity among respondents^23^.

A non-random, purposive sampling will be used to identify targets participants through a literature scan of MEDLINE. In order to preserve anonymity of participants, the complete search strategy is not reported. Eligible participants will be authors of at least 2 publications of any study design concerning PFD among athletes. After this phase, a set of unique authors’ names and contact information will be extracted. Then, all eligible authors will be invited by an e-mail of the first author (SG) for Delphi participation together with the research team. The mail will include a brief note underlining a) the aim of the study, b) contact name and address of the first author, c) data handling, d) privacy policy, e) informed consent, f) instructions for the completion of the survey and g) the related link invitation. Participation will be on a voluntary basis and no incentives will be offered.

Data will be downloaded and stored in an encrypted computer, and only the first author will be access to the information during all stages of the study^24^. Participants will be ensured that their identities would not be disclosed. All personal data will be de-identified (name and e-mail address) to maintain confidentiality and data protection^24^. In the online software SurveyMonkey (SurveyMonkey, Palo Alto, CA), participants will be able to review or change responses using a back button, before submitting their answers.

Two Delphi rounds will be run. The first round is planned in February 2022, the second in April 2022. Before invitation, the content of each round will be pilot tested.

Selected participants will be invited to participate in both rounds, unless they explicitly indicate that they do not wish to participate. During each round, 1 e-mail reminder will be sent to every participant. Participants will be asked about sociodemographic (e.g., nationality, age, sex) and professional characteristics (e.g., educational background, current field of work, current role, experience and number of studies on the topic).

The proposals of items will be presented in the Delphi survey as closed questions in which participants could score the endorsement of each item for inclusion in the screening tool on a 5-point Likert scale ranging from “Strongly disagree/Absolutely no” to “Strongly agree/Absolutely yes” (e.g. *Strongly agree to include the item in the screening tool for referral*) and give reasons for their answers.

Because Delphi studies rely on reaching a consensus, no sample size calculation will be calculated. A consensus will be set a priori at 67% of total number of participants (dis)agreeing with a proposal (ie, “Strongly (dis)agree” and “(Dis)Agree” answers) will be pooled together.

Consistency of results will be assessed by separately calculating proportions of each stakeholder group (i.e, researchers, clinicians).

### Delphi round 1

Initial general statements regarding the use and the importance of screening tools in the field will be incorporated in the survey.

In addition, variables and risk factors will be presented as items and participants will be asked whether they agree or disagree with the endorsement of each item for inclusion in the screening tool. Subsequently, criteria for referral will be decided by the participants.

Finally, two open questions will be asked for additional items and for generic feedback on the Delphi.

### Delphi round 2

We will present, summarize, and send the results of Round 1 to the participants, including their own ratings and comments. Items without a consensus will be presented again for voting only if they had at least 50% of participants in favor of the endorsement or if any substantial remark favored their endorsement. In case of no consensus, all potential items will be presented again for rating. Additional items will be added in this round, based on first-round participant suggestions.The round will conclude with an open question.

## Results from the Delphi survey

Item scores will be summarized as appropriate (eg, frequency and proportions) accompanied by a narrative summary of findings, comments, and suggestions.

### Screening tool for referral

The research team will participate in a focus group to discuss all the results. Once approval will be obtained from all the members, the items will be included in the screening tool and will be considered ready for reporting.

## CONCLUSIONS

PFD is an under-recognized and under-treated condition in female athletes^25^.

A screening tool will be developed to aid clinicians generally non-specialized in pelvic floor health (musculoskeletal/sport physiotherapist, medical doctors specialized in sports medicine) referring female athletes to a PFD expert such as a uro-gynecologist and a pelvic floor/women’s health physiotherapist.

This step could represent a starting point for the recognition of potential PFD among the female athletes and therefore, the related treatment pathway. Additional prospective studies will be needed to validate the tool.

## Data Availability

All data produced in the present work are contained in the manuscript

## Acknowledgements

Not applicable.

